# First in human measurements of abscess cavity optical properties and methylene blue uptake prior to photodynamic therapy by *in vivo* diffuse reflectance spectroscopy

**DOI:** 10.1101/2023.10.16.23297088

**Authors:** Md Nafiz Hannan, Ashwani K. Sharma, Timothy M. Baran

## Abstract

**Significance:** Efficacious photodynamic therapy (PDT) of abscess cavities requires personalized treatment planning. This relies on knowledge of abscess wall optical properties, which we report for the first time in human subjects.

**Aim:** The objective was to extract optical properties and photosensitizer concentration from spatially-resolved diffuse reflectance measurements of abscess cavities prior to methylene blue (MB) PDT, as part of a Phase 1 clinical trial.

**Approach:** Diffuse reflectance spectra were collected at the abscess wall of 13 human subjects using a custom fiber-optic probe and optical spectroscopy system, before and after MB administration. A Monte Carlo lookup table was used to extract optical properties.

**Results:** Pre-MB abscess wall absorption coefficients at 665 nm were 0.15±0.1 cm^−1^ (0.03-0.36 cm^−1^) and 10.74±15.81 cm^−1^ (0.08-49.3 cm^−1^) post-MB. Reduced scattering coefficients at 665 nm were 8.45±2.37 cm^−1^ (4.8-13.2 cm^−1^) and 5.6±2.26 cm^−1^ (1.6-9.9 cm^−1^) for pre-MB and post-MB, respectively. Oxygen saturations were found to be 58.83±35.78% (5.6-100%) pre-MB and 36.29±25.1% (0.0001-76.4%) post-MB. Determined MB concentrations were 71.83±108.22 µM (0-311 µM).

**Conclusions:** We observed substantial inter-subject variation in both native wall optical properties and methylene blue uptake. This underscores the importance of making these measurements for patient-specific treatment planning.

## 1. Introduction

Abscesses, which consist of a purulent collection surrounded by a fibrous pseudo-capsule, form as a result of host immune and inflammatory response to an acute bacterial infection^1^. Without treatment, these localized pockets of infection can rupture or spread systemically, leading to high rates of morbidity and mortality^2^. Although image-guided percutaneous drainage has become the standard of care for abscesses that do not respond to antibiotics alone^3^, abscess resolution rates can be low in certain cases^4^ and complications following drainage are a persistent problem^5,6^. Furthermore, the occurrence rate of antibiotic-resistant bacteria is on the rise, with multi-drug resistant pathogens increasingly found in abscess aspirates^7,8^. Alternative treatments are therefore needed.

Photodynamic therapy (PDT), which produces an antimicrobial effect through the photochemical production of reactive oxygen species, may be a powerful adjunct to drainage in the treatment of infected abscesses. Multiple studies have shown that PDT is efficacious *in vitro* against bacterial species that are typically found in abscess cavities^7,9^. Based on these promising results, we completed a Phase 1 clinical trial studying the safety and feasibility of PDT with the photosensitizer methylene blue (MB) at the time of abscess drainage (ClinicalTrials.gov Identifier: NCT02240498). The primary clinical outcomes of this trial are reported elsewhere^8^. Here, we focus on the results of optical spectroscopy measurements made immediately prior to PDT.

PDT efficacy is largely determined by the combination of the absorbed light dose and the photosensitizer concentration^10^. The absorbed light dose is determined by the treatment laser power and the optical properties, absorption and scattering, of the target tissue. If the tissue optical properties are known, forward modeling can be used to determine the distribution of light dose within the target tissue. Given a target light dose, patient-specific treatment plans can then be generated to ensure that the entire target region receives an efficacious light dose.

This type of patient-specific PDT treatment planning has been widely applied for interstitial PDT of cancer^11^. For example, Davidson *et al* used the treatment diffuser fibers to extract optical properties of the prostate based on a diffusion model^12^. These optical properties were then used to design treatment plans that delivered a threshold dose to the prostate plus margin. Lietke *et al* used a similar approach in designing treatment plans for malignant glioma recurrences^13^. In the context of a larger body space, Dupre *et al* reported on the performance of surface contact spatially-resolved diffuse reflectance spectroscopy prior to PDT of the pleural cavity^14^. These measurements, along with real-time fluence monitoring^15^, were used to deliver uniform light doses to highly heterogeneous subjects.

While other groups have reported optical property measurements within the human abdomen^16,17^, the optical properties of human abscesses have never been measured, making accurate light dose modeling impossible. In order to remedy this, we have designed, built, and validated a spatially-resolved diffuse reflectance spectroscopy system incorporating multiple optical fibers within a 2 mm outside diameter package^18,19^. As described above, many prior applications typically rely on multiple distinct spectroscopy fibers^12,13,20^, large area of access to the treatment site^14,15^, or bulky, rigid fiber optic probes^21–23^ in order to extract patient-specific optical properties for treatment planning. In the case of abscesses, access is generally provided by a small drainage catheter and avoidance of abscess rupture is paramount. The small outside diameter (∼2 mm) and flexibility of the fiber-optic probe are therefore crucial for integration into the clinical workflow.

In this study, we performed diffuse reflectance spectroscopy immediately prior to PDT on a series of 13 subjects treated as part of the parent Phase 1 clinical trial. These spectroscopy measurements were taken both before and after methylene blue (MB) administration, in order to quantify native abscess wall optical properties and MB uptake. This represents the first report on the optical properties of human abscess cavities, as well as the retention of MB following a 10 minute incubation. These optical property measurements set the stage for patient-specific treatment planning^24^, building on our retrospective study results^25,26^. We previously reported preliminary results for the first three subjects measured in this study^27^. Here, we present the full results for the entire study cohort, including additional analyses.

## 2. Materials and Methods

### 2.1. Participants and Regulatory Approval

Spectroscopy data were collected from subjects enrolled in a Phase 1 clinical trial examining the safety and feasibility of methylene blue photodynamic therapy performed at the time of percutaneous abscess drainage (ClinicalTrials.gov Identifier: NCT02240498). The primary results of this clinical trial are reported separately^8^. Of the 18 subjects treated in this trial, optical spectroscopy data were collected from the final 13 subjects as described below. For the first 5 subjects treated, funding was not available to construct the spectroscopy system described in Section 2.2 and perform spectroscopy measurements. All study procedures were approved by the Research Subjects Review Board at the University of Rochester Medical Center, and written informed consent was provided by all subjects.

### 2.2. Diffuse Reflectance Spectroscopy System

Spatially-resolved diffuse reflectance data at the abscess wall were collected using an optical spectroscopy system that was described in detail previously^18^, as shown in Figure 1a. Briefly, this system consists of an optical switch (FSM 1×2, Piezosystem Jena, Inc., Hopedale, MA) that routes either broadband white light (HL-2000-HP-FHSA, Ocean Optics, Inc., Largo, FL) or laser light at 640 nm (OBIS FP 1193841, Coherent, Inc., Santa Clara, CA) to the source fiber on the optical probe displayed in Figures 1b and 1c. Light transmitted by each of the 8 detector fibers is sequentially routed to a spectrometer (QE Pro, Ocean Optics) for detection via another optical switch (FSM 1×8, Piezosystem Jena, Inc.). Detected spectra were corrected for dark background and integration time, and were each divided by a corresponding calibration measurement made using an integrating sphere (3P-GPS-020-SF, Labsphere, Inc., North Sutton, NH) incorporated into the system housing. Details of the calibration procedure are reported elsewhere^18^. While we have examined the fluorescence data collected by this system pre-clinically^19^, the focus of the current study is the analysis of diffuse reflectance data.

**Figure 1:**
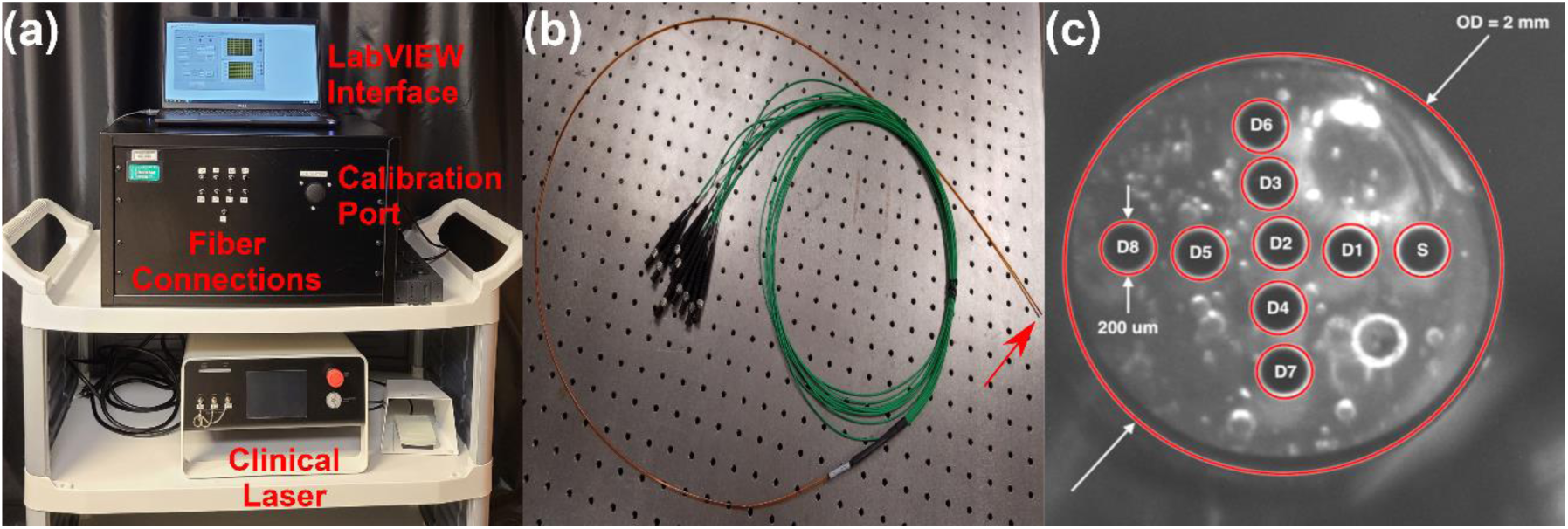
(a) Image of spectroscopy system showing fiber connections for source and detector fibers, port for integrating sphere calibration, LabVIEW interface, and the laser used for clinical PDT. (b) Image of fiber optic probe, with distal tip indicated by red arrow. (c) Image of fiber optic probe distal face showing the 200 µm diameter fibers used for delivery and detection of light. The source fiber is labeled “S,” detector fibers are labeled “D,” and outside diameter is abbreviated as “OD.”

The fiber optic probe used for delivery of incident light and detection of diffuse reflectance (Pioneer Optics Company, Bloomfield, CT) is shown in Figure 1b. This flexible probe has an outside diameter of 2 mm, in order to allow it to be inserted through the standard of care drainage catheter used for percutaneous abscess drainage, as described in Section 2.3. In order to determine the source-detector separations necessary for determination of optical properties from measured spectra, images were captured of the probe face using a stereomicroscope with a large working distance (SMZ1500, Nikon Instruments, Inc., Melville, NY). These images were captured with and without a United States Air Force target (USAF 1951 1X, Edmund Optics, Inc., Barrington, NJ) in the frame to translate pixel coordinates to distance. These source-detector separations ranged from 320-1310 µM, as shown in Figure 1c.

As described by Bridger *et al*, these source-detector separations were used to generate a Monte Carlo lookup table for extraction of optical properties from measured spectra^18^. Whereas this prior publication used a lower resolution color camera to determine fiber positions, the current results utilize the more precise positions determined by microscopic evaluation. Otherwise, the procedure was identical to the previously reported multispectral fitting method. Based on visual inspection of measured spectra, absorption spectra were fit as a superposition of oxy- and deoxy-hemoglobin, MB monomer, and MB dimer absorption spectra. MB dimer absorption was included as MB dimerizes at high concentrations and there is an affiliated change in the absorption spectrum^28^. Reduced scattering spectra were assumed to follow a power-law relationship of the form 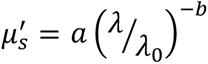, where *λ* corresponds to wavelength in nm and *λ_0_* is a normalization wavelength set to 665nm.

### 2.3. Clinical Data Collection

All subjects first had their abscess drained using standard of care image-guided percutaneous drainage, as described in the main trial report^8^. This procedure utilized CT guidance for 16 subjects and ultrasound guidance for 2 subjects. Immediately following drainage, the disinfected optical probe was removed from the bag and the proximal fiber ends connected to the spectroscopy system. The distal end was then held in place at the entrance port of the system integrating sphere by the interventional radiologist. Calibration spectra were captured at each detector fiber for both broadband and laser illumination, with an integration time of 100 ms for broadband illumination and 50 ms for laser illumination. These calibration spectra were used to correct patient data as described in Section 2.2.

In order to measure patient optical properties, the interventional radiologist advanced the optical probe through the standard of care drainage catheter until it made gentle contact with the abscess wall. Surface contact was insured through tactile feedback by the study doctor, and by monitoring the signal detected at the closest detector fiber in real time. As described by Chen *et al*^29^, detected reflectance increases as the probe approaches the surface before rapidly decreasing when surface contact is made. This is due to the transition from specular to diffuse reflectance.

Once surface contact was made, diffuse reflectance and fluorescence spectra were captured at each detector fiber sequentially. For each fiber, an initial integration time of 50 ms was used. Assuming a linear relationship between integration time and detected signal, the maximum magnitude of the detected signal was used to calculate the integration time that would result in 50% of the dynamic range of the spectrometer being utilized. This was chosen to compromise between signal to noise ratio and measurement time. A measurement was then performed at this calculated integration time. Dark spectra were also collected at this point as described in Section 2.2 using the same integration times by disabling the source switch. If the captured spectra were not of sufficient quality, the probe was withdrawn and replaced by the study doctor, and spectroscopy was repeated. This measurement was meant to capture native abscess wall optical properties prior to MB infusion, and is referred to as the “pre-MB” measurement elsewhere.

Following this, 0.1% methylene blue (BPI Labc, LLC, Largo, FL) was infused into the abscess cavity and incubated for 10 minutes to allow for bacterial uptake. MB was then aspirated and the cavity was flushed with sterile saline. The optical probe was then re-inserted through the drainage catheter into gentle contact with the abscess wall, and measurements were repeated as described above. This measurement was meant to capture the uptake of methylene blue, and is referred to as the “post-MB” measurement throughout.

After the completion of optical spectroscopy measurements, the optical probe was removed and photodynamic therapy was performed as described in the main clinical trial report^8^. This consisted of infusion of a lipid emulsion to scatter treatment light through the irregular abscess cavity, insertion of a sterile optical fiber, and delivery of laser light at 665 nm.

### 2.4. Data Processing

As described above, collected spectra were corrected for dark background and integration time, and divided by corresponding spectra collected at the entrance port of the system integrating sphere. Optical property spectra were extracted using the Monte Carlo lookup table approach described in Section 2.2. Whereas pre-clinical validation of the system used all eight detector fibers, high-quality data could not be collected at all detector fibers for all subjects in clinical measurements. This was mainly due to poor surface contact between specific detector fibers and the abscess wall, even after repositioning of the optical probe. Additionally, for subjects with high MB uptake, spectra collected at larger source-detector separations did not have sufficient signal to noise ratio for analysis due to major attenuation by methylene blue absorption. In cases where spectra at individual detectors were not usable, these detected fibers were excluded from the optical property inversion. As we have shown previously, utilization of fewer source-detector separations does not significantly impact the accuracy of optical property recovery, as long as data from at least 4-5 detector fibers are included^30,31^. This was achievable for all but the first subject, resulting in 13 pre-MB measurements and 12 matched post-MB measurements.

### 2.5. Statistical Analysis

Continuous values are summarized across subjects as mean ± standard deviation, while categorical values are summarized as proportions with 95% confidence intervals. Demographic information was compared between the 13 subjects that received optical spectroscopy and the five treated prior to construction of the optical spectroscopy system using the Mann-Whitney test for continuous variables and Fisher’s exact test for categorical values. Extracted optical properties at 665 nm and SO_2_ were compared between pre- and post-MB measurements using the Wilcoxon signed-rank test. Correlation between MB concentration and change in SO_2_ was calculated using the Spearman correlation coefficient. Optical properties were compared between abscess locations using the Kruskal-Wallis test, with Dunn’s test for pairwise comparisons. A p value less than 0.05 was considered significant, and all analyses were performed in GraphPad PRISM (v6, GraphPad Software, Inc., San Diego, CA) and MATLAB (R2022b, The Mathworks, Inc., Natick, MA).

## 3. Results

### 3.1. Subject demographics

Subject demographics for all subjects that received PDT, as well as separated by whether optical spectroscopy measurements were made, are included in Table 1. As can be seen, there were no significant differences in demographics between subjects that did and did not receive optical spectroscopy. For the 13 subjects described in this report, 38.5% were male and the average age was 56.9±20.3 years.

**Table 1.** Subject demographics for overall sample, and separated by whether spectroscopy was performed. P values are the results of the Mann-Whitney test for continuous variables and Fisher’s exact test for categorical variables, comparing those for which optical spectroscopy was and was not performed.

|  | Overall Sample (n=18) | Optical Spectroscopy Performed (n=13) | Optical Spectroscopy Not Performed (n=5) | P value |
| --- | --- | --- | --- | --- |
| Age | 60.1±18.3 | 56.9±20.3 | 68.6±7.6 | 0.20 |
| Sex (% Male) | 44.4% (21.5-69.2%) | 38.5% (13.9-68.4%) | 60.0% (14.7-94.7%) | 0.61 |
| BMI | 29.3±5.9 | 30.0±6.3 | 25.4±6.1 | 0.39 |
| Abscess Volume (mL) | 36.8±40.6 | 25.2±22.0 | 67.0±63.0 | 0.18 |

### 3.2. Representative data from two subjects

Representative diffuse reflectance spectra for two subjects are shown in Figure 2, with fully corrected data shown as open circles and best fit spectra shown as solid lines. As can be seen in Figures 2a and 2c, pre-MB data were accurately fit with a super-position of oxy- and deoxy-hemoglobin absorption and power-law reduced scattering. Post-MB data, as shown in Figures 2b and 2d, were well fit by the addition of methylene blue monomer and dimer spectra.

**Figure 2:**
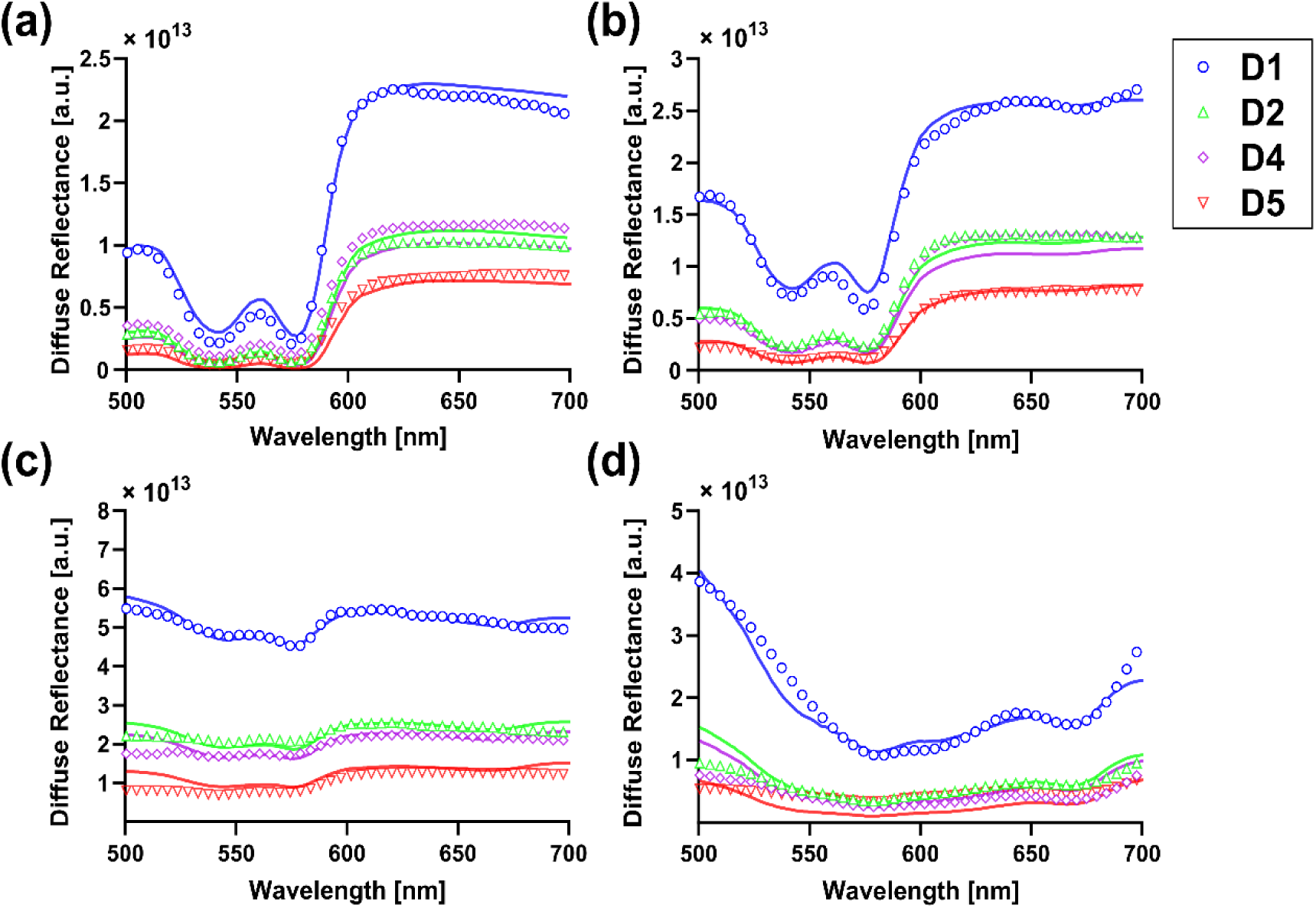
Corrected diffuse reflectance data (open circles) and fits (solid line) for detector fibers 1, 2, 4, and 5 for Subject 3 (a) pre-MB and (b) post-MB, and Subject 8 (c) pre-MB and (d) post-MB. Colors and symbols indicate different detector fibers.

These fits were used to extract the optical property spectra shown in Figure 3. As can be seen, there was variability between subjects in both pre-MB optical properties and MB uptake. Figures 3a and 3c demonstrate large differences in absorption spectra between subjects for both pre-MB and post-MB conditions. In both subjects, oxy- and deoxy-hemoglobin were the main absorbers.

**Figure 3:**
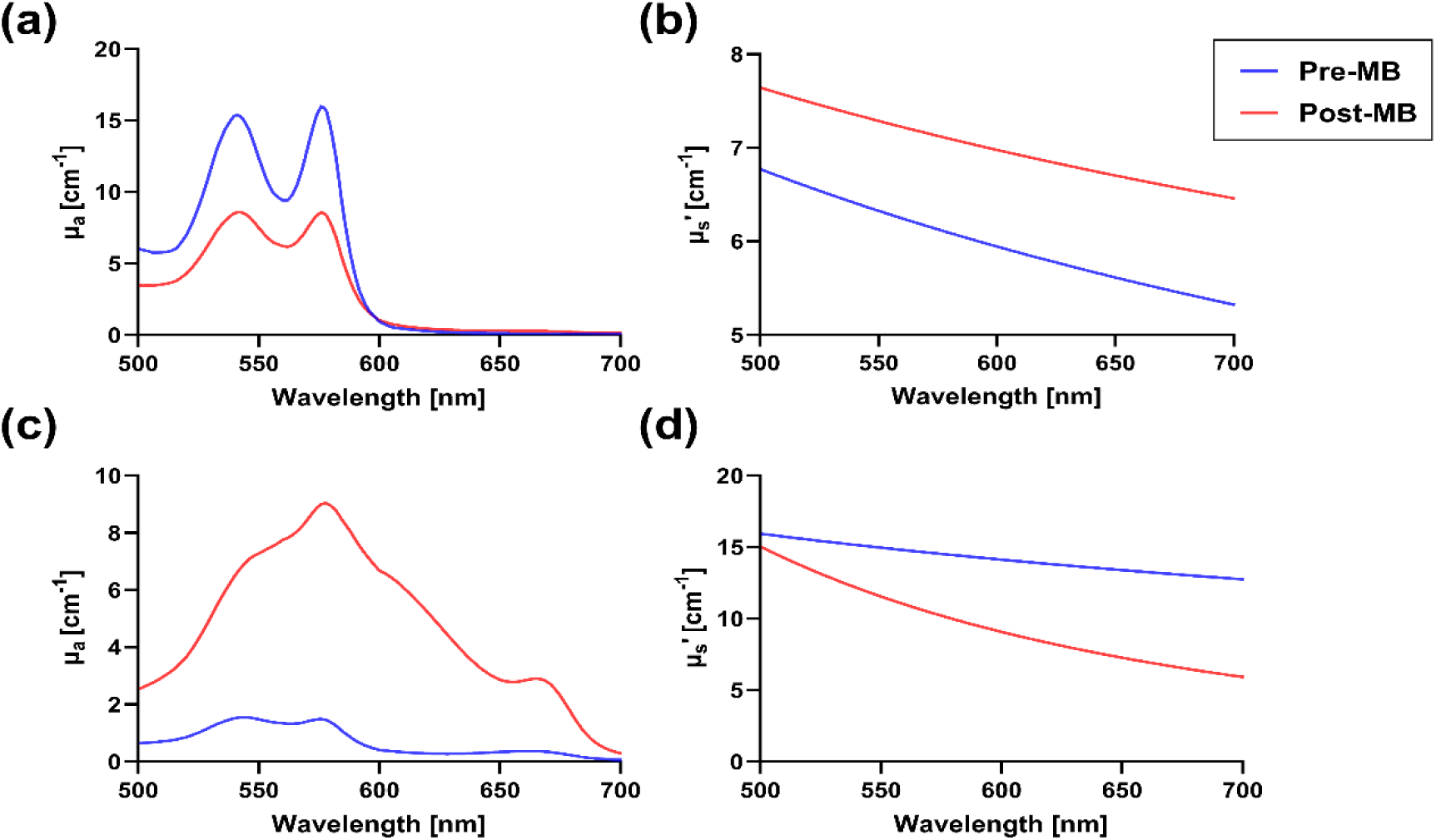
Recovered (a) absorption (µ_a_) and (b) reduced scattering (µ_s_’) spectra for subject 3 shown in Figures 2a and 2b. Recovered (c) µ_a_ and (d) µ_s_’ spectra for subject 8 shown in Figures 2c and 2d. Blue and red lines represent pre-MB and post-MB conditions, respectively.

In the post-MB condition for subject 3, oxy- and deoxy-hemoglobin remained the dominant absorbers. For subject 8, methylene blue monomer and dimer dominated the post-MB absorption spectra, with some contributions from oxy- and deoxy-hemoglobin. Figures 3b and 3d illustrate notable differences in reduced scattering spectra between subjects for pre-MB and post-MB conditions.

### 3.3. Native abscess wall optical properties

Native, pre-MB abscess wall optical properties at 665 nm across subjects are tabulated in Table 2. Substantial inter-patient variability in native abscess-wall optical properties and oxygen saturation were observed (Figure 4). Means and standard deviations for absorption coefficient at 665 nm (µ_a,665_) were 0.15 ± 0.1 cm^−1^ (range: 0.03-0.36 cm^−1^), and 8.45 ± 2.37 cm^−1^ (range: 4.8-13.2 cm^−1^) for reduced scattering coefficient at 665 nm (µ_s,665_’). Oxygen saturation (SO_2_) was found to be 58.83 ± 35.78% (range: 5.6-100%).

**Table 2.**
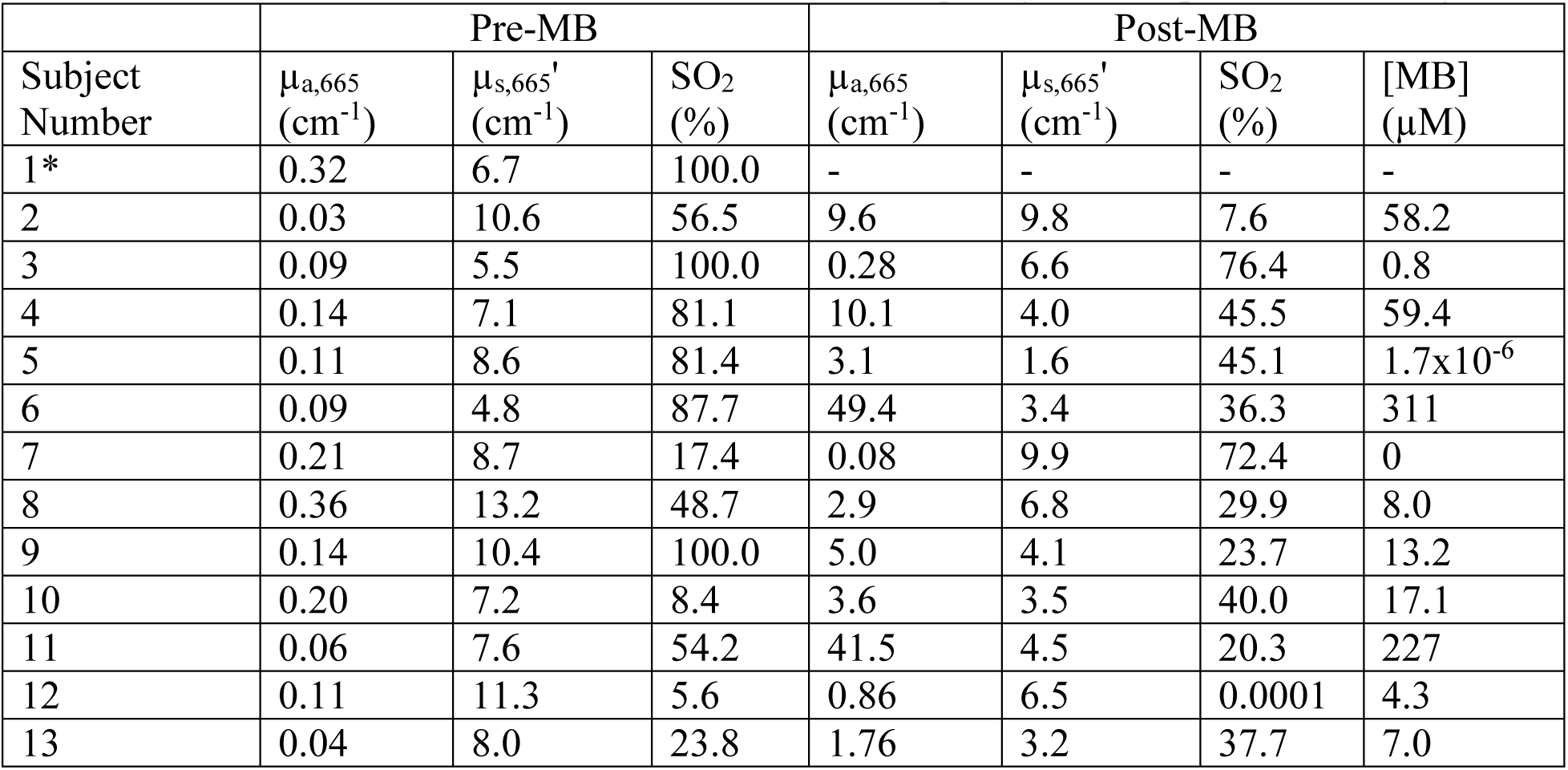
Extracted optical properties at 665 nm and oxygen saturation for each subject, before and after addition of methylene blue (MB). MB concentration ([MB]) is also included for post-MB measurements. *Post-MB data were not of sufficient quality for interpretation for subject 1.

| Subject Number | Pre-MB |  |  | Post-MB |  |  |  |
| --- | --- | --- | --- | --- | --- | --- | --- |
| | $\mu_{a,665}$ (cm<sup>-1</sup>) | $\mu_{s,665}'$ (cm<sup>-1</sup>) | SO<sub>2</sub> (%) | $\mu_{a,665}$ (cm<sup>-1</sup>) | $\mu_{s,665}'$ (cm<sup>-1</sup>) | SO<sub>2</sub> (%) | [MB] (μM) |
| 1* | 0.32 | 6.7 | 100.0 | - | - | - | - |
| 2 | 0.03 | 10.6 | 56.5 | 9.6 | 9.8 | 7.6 | 58.2 |
| 3 | 0.09 | 5.5 | 100.0 | 0.28 | 6.6 | 76.4 | 0.8 |
| 4 | 0.14 | 7.1 | 81.1 | 10.1 | 4.0 | 45.5 | 59.4 |
| 5 | 0.11 | 8.6 | 81.4 | 3.1 | 1.6 | 45.1 | 1.7x10<sup>-6</sup> |
| 6 | 0.09 | 4.8 | 87.7 | 49.4 | 3.4 | 36.3 | 311 |
| 7 | 0.21 | 8.7 | 17.4 | 0.08 | 9.9 | 72.4 | 0 |
| 8 | 0.36 | 13.2 | 48.7 | 2.9 | 6.8 | 29.9 | 8.0 |
| 9 | 0.14 | 10.4 | 100.0 | 5.0 | 4.1 | 23.7 | 13.2 |
| 10 | 0.20 | 7.2 | 8.4 | 3.6 | 3.5 | 40.0 | 17.1 |
| 11 | 0.06 | 7.6 | 54.2 | 41.5 | 4.5 | 20.3 | 227 |
| 12 | 0.11 | 11.3 | 5.6 | 0.86 | 6.5 | 0.0001 | 4.3 |
| 13 | 0.04 | 8.0 | 23.8 | 1.76 | 3.2 | 37.7 | 7.0 |

**Figure 4:**
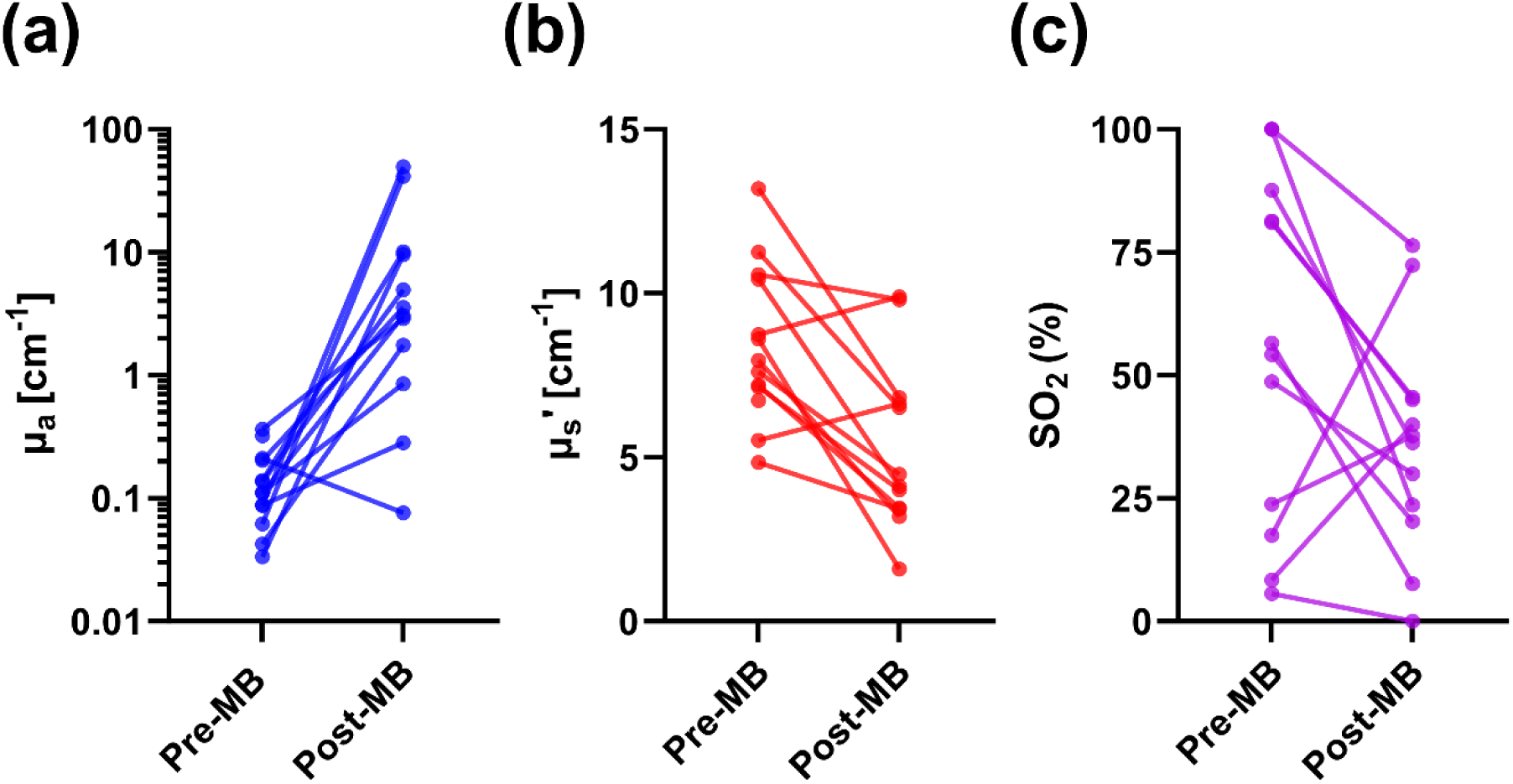
Extracted (a) absorption coefficients (µ_a_) at 665 nm, (b) reduced scattering coefficients (µ_s_’) at 665 nm, and (c) oxygen saturation (SO_2_) for all subjects for pre-MB and post-MB conditions. Note that (a) is on a log scale, while (b) and (c) are on linear scales.

### 3.4. Methylene blue uptake and effects on extracted optical properties

Measured MB concentrations across subjects are tabulated in Table 2. Mean and standard deviation for this was 71.83 ± 108.22 µM (range: 0-311 µM), which reveals substantial inter-patient variation. As expected, there was a significant increase in µ_a_ at 665 nm for post-MB measurements (p=0.001), due to the presence of methylene blue absorption (Figure 4a). There was also an apparent decrease in µ_s_’ at 665 nm (p=0.005, Figure 4b). While there was a decrease in SO_2_ from pre- to post-MB values (median reduction = 28.8%, Figure 4c), this difference was not significant (p=0.11). Additionally, this change in SO_2_ from pre- to post-MB measurement was not significantly correlated with measured MB concentration (ρ=0.49, p=0.11).

As the fiber optic probe was removed and reinserted between the pre- and post-MB data collection, some of this variability may also represent small differences in the exact placement of the probe face.

### 3.5. Differences based on abscess location or bacteria present

For the 13 abscesses that were quantified optically, four were located in the pelvis, five were associated with the colon, and four were found in other locations. While sample sizes were small, we performed preliminary analysis to examine regional differences in optical properties. None of the quantities reported in Table 2 were found to be significantly different between these rough location groupings.

Of the bacterial species found in abscesses measured, the most common was *Escherichia coli* (*E. coli*, n=3). Other abscess cavities were found to contain a variety of other species. While abscess containing *E. coli* tended to have higher MB uptake than those with other bacterial species (97.7±114.7 µM vs. 14.8±22.7 µM), this difference was not statistically significant (p=0.17). Four of the 13 abscess measured were found to contain antibiotic-resistant bacterial strains. MB uptake was not different between strains that were resistant or susceptible to antibiotics (p=0.63), indicating that PDT susceptibility in antibiotic-resistant strains would not be limited by MB concentration.

## 4. Discussion

We performed clinical spatially-resolved diffuse reflectance spectroscopy of human abscess cavities prior to PDT, representing the first time that the optical properties of human abscesses have been quantified. There was substantial variation in the native abscess wall optical properties prior to methylene blue administration, particularly in absorption at 665 nm and SO_2_. Uptake of methylene blue, as measured by post-MB diffuse reflectance, varied widely between subjects. Infusion of MB appeared to reduce SO_2_ measured at the abscess wall, with minimal effects on tissue scattering. There were no significant differences based on abscess location or bacteria present.

This represents the first study that measured the optical properties of human abscesses. As such, direct comparisons to perfectly analogous results are impossible. However, other investigators have reported optical properties measured within nearby anatomy in the abdomen. Perhaps the most thorough of these is Morales *et al*, where diffuse reflectance measurements were made with a surface-contact probe throughout the peritoneal cavity^16^. This included measurements at the surface of multiple organs, the chest wall, and skin. Pre-MB absorption and reduced scattering coefficients determined in this study fall within the ranges reported by these investigators. Wang *et al* also measured optical properties within the peritoneal cavity prior to PDT^32^. Although their results are provided at 630 nm rather than 665 nm, our reported values are similar to those reported for the peritoneum, small bowel, and large bowel. We have also previously measured the optical properties of excised human kidneys^30^, finding that absorption and reduced scattering at 665 nm ranged from 0.3-1.7 cm^−1^ and 12.5-39.4 cm^−1^, respectively. These ranges are again similar to what we find in the present study. Abscesses are not routinely surgically excised, as this significantly increases morbidity and mortality^33^. Capture of biopsy samples is also not possible, as this could rupture the cavity and lead to sepsis. We therefore do not have patient tissue available to perform optical property quantification via other means. Based on these other reports, however, it appears that our measurements are in reasonable concordance with similar abdominal tissue.

Oxygen saturation (SO_2_), as determined by the relative concentrations of oxy- and deoxy-hemoglobin, was found to decrease from the pre- to post-MB measurements (55.4±35.1% vs. 36.2±22.7%) and this decrease was correlated with measured MB concentration (ρ=0.49). However, this change was not statistically significant (p=0.11). This suggests that the infusion of a high MB concentration, even after flushing the cavity with sterile saline, may be related to a reduction in SO_2_. One potential explanation is localized oxidation of hemoglobin to methomoglobin, which cannot bind oxygen^34^, at high MB concentrations^35^. As we rely on the balance between oxy- and deoxy-hemoglobin to determine SO_2_, any changes in the ability of hemoglobin to bind oxygen would be incorrectly interpreted as a reduction in the availability of oxygen. Other studies have shown that systemic administration of MB can also result in artificially low SO_2_ readings on standard pulse oximetry^36–38^. Importantly, we did not observe changes in SO_2_ with standard of care pulse oximetry on the finger, indicating that these changes are localized to the region of MB administration.

As described in Section 2.2, we use a multispectral fitting procedure that includes absorption contributions from hemoglobin (oxy- and deoxy-) and methylene blue (monomer and dimer). For the range of MB concentrations we observed here, the inclusion of MB dimer absorption was of particular importance for post-MB measurements. MB forms dimers in aqueous solution, with the dimer fraction correlated with MB concentration^39^. The absorption of this dimer is shifted from the monomer peak of 664 nm to approximately 600 nm. In addition to more accurate fitting in the spectral region around 600 nm, the inclusion of MB dimer absorption also had a profound effect on the accuracy of SO_2_ determination. An example of this is shown in Figure 5. Without the MB dimer in the fit the spectral shape is not fit well and an erroneously low SO_2_ value is returned. When the MB dimer is properly included (see Figure 2d), the fit is of much higher quality and the correct value of SO_2_ is found. For the example shown in Figure 5, the SO_2_ value increased from 0% to 29.9% by inclusion of the dimer. Across all subjects, if fits were performed with the MB dimer absorption included, the determined SO_2_ post-MB was significantly increased (36.2±22.7% vs. 25.9±27.3%, p=0.027). This highlights the importance of incorporating all relevant absorbers into spectral fitting.

**Figure 5:**
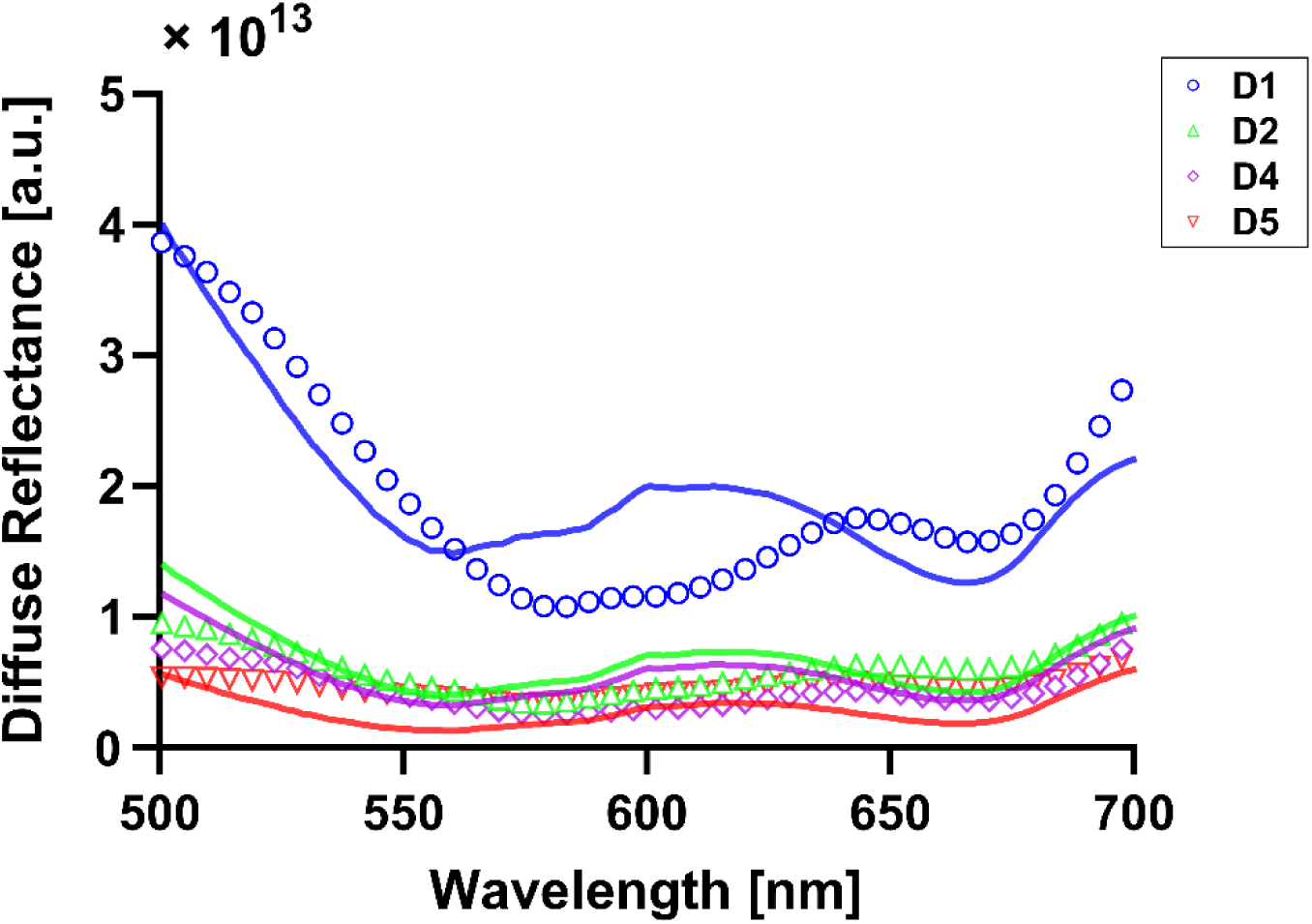
Effect of excluding MB dimer from fitting for the same subject shown in Figure 2d. Colors and symbols represent different detector fibers.

As mentioned previously, MB dimer formation generally occurs at high MB concentration. Taken together with the observation of negative correlation between MB uptake and SO_2_, this motivates reduction of MB concentration in future studies. This is further supported by pre-clinical studies, where MB concentration is typically much lower than the 1 mg/mL concentration used here. For example, Haidaris *et al* showed efficacious PDT against multiple bacterial species isolated from patient abscess aspirates at a MB concentration of 300 µg/mL^7^. In an *in vivo* study in cows, Sellera *et al* showed that a MB concentration of 100 µg/mL (0.01%) was efficacious in treating bovine skin infection when combined with red LED illumination^40^. This 0.01% (100 µg/mL) MB concentration was also shown to be efficacious in controlling *Streptococcus mutans* biofilms^41^. For clinical studies of MB-PDT, concentrations ranging from 25-100 µg/mL have typically been reported^42–44^, with some investigators using higher concentrations for topical application^45^. However, these studies using higher concentrations typically employ much higher fluence rates^46^, which are not applicable in the case of abscesses.

The wide variability in extracted optical properties, particularly after the addition of MB, provides a strong motivation for the performance of patient-specific treatment planning in future PDT of abscesses. In prior retrospective studies, we have shown the importance of this for improving eligibility for PDT^25,26^. Further, the optical properties reported here have been used to generate retrospective treatment plans for the 13 subjects that received both PDT and spectroscopy^24^. In particular, the effects of abscess wall absorption have a pronounced effect on generated treatment plans. Along with the evidence discussed above, this motivates a reduction in methylene blue concentration in future studies. While prior studies in oncological PDT have employed patient-specific treatment planning^11–13^, none of these studies have directly compared patient-specific treatment plans to “fixed dose” plans that do not incorporate patient-specific optical properties. This motivates a direct comparison between a “fixed dose” case and patient-specific treatment planning in upcoming clinical trials.

We acknowledge some limitations in the present study. These findings are from a relatively small (n=13) group of subjects treated with PDT at the time of their abscess drainage, so results could be vulnerable to bias. While a large number of the abscesses measured presented in the pelvis (n=4), many were located in other anatomical locations. There is therefore insufficient sample size to make conclusive statements about regional differences in optical properties. Additionally, extraction of optical properties required the fiber optic probe to make surface contact with the abscess wall. While we used both tactile and optical feedback to verify this, it was apparent in some cases that certain detector fibers were not in contact with the abscess wall. This meant that extraction of optical properties in these cases was based upon a smaller number of detector fibers, which could reduce precision. This motivates the design of alternate probe geometries for future studies. Finally, while measurements were made before and after the addition of MB, no measurements were made after PDT. This has been done by other groups for PDT of cancer^16,47^, in order to quantify photosensitizer bleaching and PDT-induced changes in optical properties.

## Data Availability

All data produced in the present study are available upon reasonable request to the authors

## Disclosures

The authors have no conflicts of interest to disclose.

### Code, Data, and Materials

The data that support the findings of this article are not publicly available due to privacy concerns, as research subjects did not consent to use of their data beyond the purposes of this study.

## Acknowledgments

The authors would like to thank Dr. Joan Adamo, Laurie Christensen, Erica Longbine, and Maria Favella for regulatory support. This work was funded by grant EB029921 from the National Institutes of Health.

